# Single Injection Nerve Blocks Plus IV Lidocaine Infusions as an Alternative to Continuous Nerve Blocks for Perioperative Pain Management: A retrospective review

**DOI:** 10.1101/2022.02.22.22271279

**Authors:** Dmitriy Gromov, Jeremy Kearns, Jacques E Chelly

## Abstract

**Purpose:** Continuous nerve blocks (CNBs) and intravenous lidocaine infusions (IV Lido) represent an effective approach to perioperative pain management. We hypothesized that a single injection nerve block (SNB) plus intravenous lidocaine infusion (IV Lido) would be as effective as CNBs. Furthermore, since recently, the use of facial plane blocks are increasingly advocated, we compared CNBs vs SNBs plus IV Lido in patients undergoing erector spinae plane (ESP) and quadratus lumborum (QL) blocks for video assisted thoracic surgery (VATS) and abdominal/retroperitoneal surgeries, respectively.

**Patients and Method:** Using our IRB approved registry (PRO10120146), we retrospectively reviewed the electronic record of 105 patients, including 51 patients who underwent VATS and received either single injection erector plane block (SESPB) plus IV lido or continuous erector spinae plane block (CESPB), and 54 patients who underwent major abdominal surgery and received either single injection quadratus lumborum block (SQLB) plus IV Lido or continuous quadratus lumborum block (CQLB). Demographics, verbal pain scores (0-10), and opioid consumption (morphine intravenous equivalent; MIVE), all in the context of the same multimodal approach (acetaminophen, ketamine, dexmedetomidine, and ketorolac) were collected at 24, 48, and 72 hours after surgery. Alpha was set to 0.05.

**Results:** SNBs plus IV Lido were as effective as CNBs regarding pain control and total MIVE at 24, 48, or 72 hours after surgery. Subgroup analysis indicated similar findings were observed in patients who underwent VATS or major abdominal surgery.

**Conclusions:** This analysis suggests that SNBs plus IV Lido are as effective as CNBs for perioperative pain management when ESP or QL blocks are used for VATS or major abdominal surgery, respectively.

## Introduction

A continuous nerve block (CNB) is an established approach to provide effective perioperative pain management. One of the recognized benefits of CNBs is that, compared to a single injection nerve block (SNB), CNBs can significantly prolong the duration of analgesia.^1,2^ Despite the prolonged duration of analgesia, there are disadvantages and complications associated with CNBs. Disadvantages include frequent dislodgment of the perineural catheter, leaking of the local anesthetic solution, exposure of the perineural catheters related to the patients’ movements. Complications include major hematoma, infection, and failure to provide effective analgesia because of the initial misplacement or secondary dislodgment of the catheter^3^ and limiting patients’ mobility. Furthermore, limitations exist to the use of CNB in patients receiving anticoagulants. Last, in 2009, Centers for Medicare and Medicaid Services (CMS) unbundled the payment for a CNB from the payment for the follow up visit, thus reducing reimbursement. This led to a significant decrease in the number of CNBs being performed, especially in a busy practice, as placement of a CNB takes significantly more time than placement of a SNB and no additional payment was provided.^4^ Developing an alternative approach to CNBs to allow provision of prolonged perioperative analgesia without CNB limitations is of major value.

For several years, our standard protocols to manage perioperative pain for a wedge resection and laparoscopic abdominal surgery included the use of an SNB combined with a continuous infusion of lidocaine at a rate of 50 mg/hr for 48 hrs or a total dose of 2,400 mg. This technique has been used in our institution in more than 1,000 patients and has been proven to be effective and safe in the context of a multimodal approach to perioperative pain management. In addition, lidocaine infusions have been used safely in abdominal, thoracic, and urologic procedures. A number of authors have confirmed the safety of the postoperative infusion of lidocaine in doses ranging from 3,200mg to 6,000mg over a 48 hour period. ^5-19^

In the past few years, the use of interfacial plane blocks has been more and more advocated,^20^ including erector spinae plane blocks (ESPB) for thoracic surgery,^21-25^ pectoralis and serratus blocks for breast surgery^26-29^ and quadratus lumborum (QL) blocks for abdominal surgery,^30^

Intravenous lidocaine (IV lido) infusions have been demonstrated to be an effective technique for postoperative analgesia in several different types of surgery, including abdominal,^31-34^ thoracic,^35,36^ and spine.^37,38^ We hypothesized that SNBs followed by IV lido would be as effective as CNBs. More specifically, single injection erector spinae plane block (SESPB) plus IV Lido would be as effective as continuous erector spinae plane block (CESPB) for video assisted thoracic surgery (VATS), and single injection quadratus lumborum block (SQLB) plus IV Lido would be as effective as continuous quadratus lumborum block (CQLB) for abdominal/retroperitoneal surgery.

## Material and methods

### Methods

Using our IRB approved registry (PRO10120146), we retrospectively reviewed the electronic records of 105 patients who received regional anesthesia and multimodal pain control from an acute pain service and underwent either unilateral video assisted thoracic surgery (VATS) or major abdominal surgery (exploratory laparotomy, gastrectomy, hyperthermic intraperitoneal chemotherapy (HIPEC), and cysto-prostatectomy surgery). Each patient included in this analysis underwent surgery using the same enhanced recovery after surgery (ERAS) protocol. For the general anesthetic, the ERAS protocol includes an induction with IV propofol and rocuronium, followed by maintenance anesthesia consisting of 1) propofol, dexmedetomidine 0.2-0.5 mcg/kg/hr, and ketamine 0.2-1 mg/kg/hr IV infusions, 2) intraoperative magnesium 2-4g IV, and 3) rocuronium boluses for muscle relaxation to maintain a train of four (TOF) 2/4 twitches. Opioid medications were strictly avoided intraoperatively. The perioperative pain protocol included acetaminophen 1g PO preoperatively, postoperatively acetaminophen 1g IV or PO every 6 hours according to each patient’s ability to tolerate oral medication, ketamine 5-10 mg/hr IV infusion, and ketorolac 15 mg IV every 8 hours for 72 hours starting in the PACU. In addition, patients who underwent VATS received a dexmedetomidine 0.2 mcg/kg/hr IV infusion for the first 24 hours of recovery in the ICU. All patients were eligible to receive postoperative opioid medications on an as needed basis (oxycodone PO, hydromorphone IV). No patients included in this study received opioid based patient-controlled analgesia (PCA).

Prior to surgery, an interfacial plane block was performed while the patient was in the pre-operative area. Each block was performed following 1) the placement of monitors to monitor blood pressure, pulse oximetry, and EKG, 2) the proper positioning of the patient, either the sitting position in the case of a unilateral ESP block for VATS or the lateral decubitus in the case of a bilateral QL2 block for abdominal surgery or a single QL2 block performed for nephrectomy, 3) verifying the patients’ consent, allergies, and the laterality of the procedure, and 4) the administration of midazolam IV 1-2 mg and/or Fentanyl IV 50-100 mcg for sedation. After proper disinfection of the skin, a curvilinear 5 MHz ultrasound probe encased in a sterile sheath and connected to a Sonosite X-Porte® (Fujifilm Sonosite, San Diego, CA) was used to determine the proper position of the nerve block needle. The landmarks for the ESP block included the transverse process of T5 vertebrae and the erector spinae plane situated immediately above the transverse process. Next, in the case of a single injection block, a 90mm 22 Gauge Touhy needle (B-Braun, Bethlehem, PA) was properly positioned under ultrasound guidance using an in-plane approach. For the QL2 block, the probe was first placed anteriorly to identif the external oblique, internal oblique, transversalis abdominis. Then, moved posteriorly to identify the transversalis fascia, and quadratus lumborum muscles. The placement of the needle in the erector spinal plan (ESP) or below the transversalis fascia (QL2) was followed by the injection of 20ml of 0.5% ropivacaine in an incremental fashion with negative aspiration between each incremental dose. In the case of a continuous block, a 20 gauge catheter was placed 5 cm beyond the tip of the 100 mm 18 gauge Touhy needle (B-Braun, Bethlehem, PA). The procedure was repeated on the contralateral side in the case of a single injection or continuous QL2 block performed for major abdominal surgery. In the recovery room, an infusion of 0.25% lidocaine at a rate of 17.5-25 mg/hr for a unilateral infusion and 35-50 mg/hr for bilateral infusion) with additional as needed boluses of 3 ml of 0.25% lidocaine (7.5 mg) every hour as needed, was begun for each continuous peripheral nerve block (2 infusions for patients who received bilateral continuous nerve blocks). These infusions continued for 3 days after surgery. A continuous infusion of IV lido 50 mg/hr was begun in the recovery room for patients who received a single injection nerve block. These infusions were continued for 2 days after surgery.

### Statistics

The primary endpoint was opioid consumption at 24 hours postoperatively (reported as morphine IV equivalents or MIE). Opioid consumption included as needed oxycodone PO and Hydromorphone IV. No patients included in this study received opioid based patient-controlled analgesia (PCA). Secondary endpoints included OPS (oral pain scores obtained from the patients using a scale of 1 (no pain) to 10 (worst possible pain) at 24, 48, and 72 hours and opioid consumption at 48 and 72 hours.

Demographic data were reported as mean ± SD, whereas opioid consumption and pain scores were reported as median (95% interval). A t-test was performed to compare demographics, and a Kruskal-Wallis test was performed to compare opioid and non-opioid consumption, pain scores, and total non-opioid consumption at each point of time. Alpha was set up at 0.05.

## Results

The overall database was comprised of 51 patients who underwent video assisted thoracic surgery (VATS) with either continuous erector spinae plane block (CESPB; n=34) or single injection erector spinae block (SESPB) plus IV lidocaine infusion (SESPB; n=17), and 54 patients who underwent major abdominal surgery (exploratory laparotomy, HIPEC, gastrectomy, cysto-prostatectomy, or partial nephrectomy) and received either continuous quadratus lumborum block (CQLB; n=31) or single injection quadratus lumborum block (SQLB; n=23) plus IV lidocaine infusion. Continuous nerve block infusions CESPB & CQLB were administered for 72 hours postoperatively. The study covered a period of 4 months from July 1 to November 1, 2019 and was conducted at The University of Pittsburgh Shadyside Hospital.

### Demographics

Overall group and sub-group comparison did not show any statistical difference with respect to age, weight, or gender (Table 1).

**Table 1.**
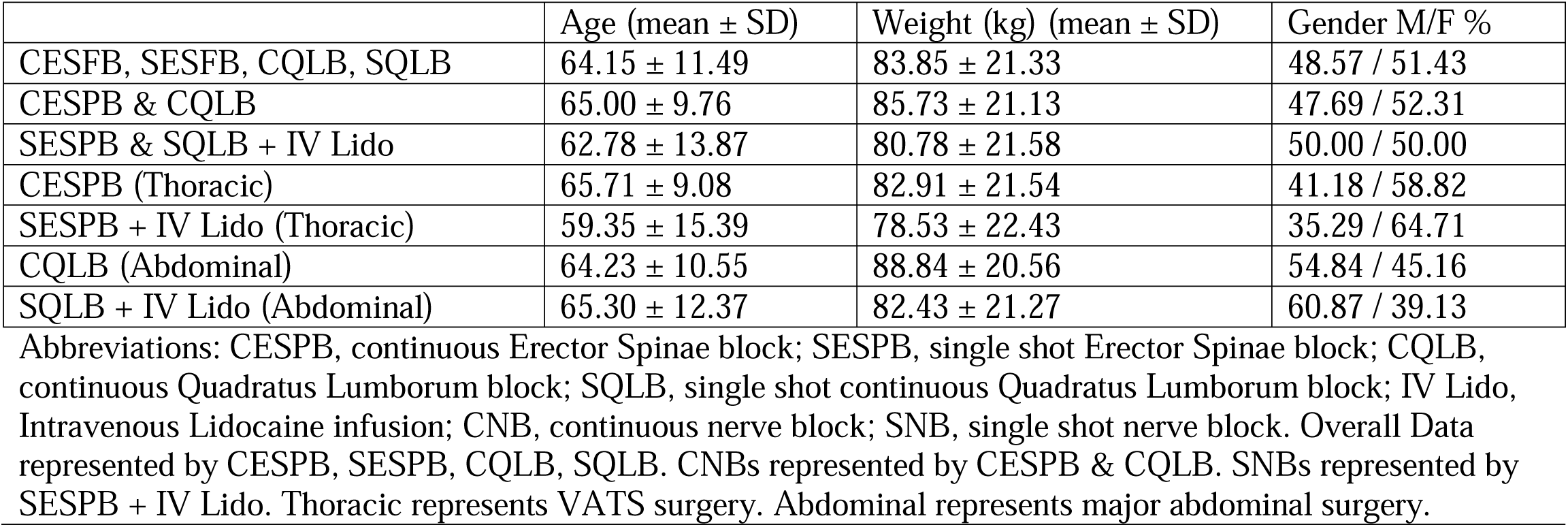
Demographic characteristics of patients (mean ± SD)

### Overall continuous nerve blocks (CNB) vs single nerve blocks (SNB) plus IV Lido

1. **Pain scores** There were no statistical differences among groups. OPS scores were as follows (Table 2):
  a. 24 hours postoperatively, CNB (5, 0.59) vs SNB + IV Lido (5, 0.75)
  b. 48 hours, CNB (4, 0.61) vs SNB + IV Lido (4, 0.94)
  c. 72 hours, CNB (4, 0.64) vs SNB + IV Lido (4, 0.94)
2. **Opioid consumption** There were no statistical differences among groups. MIE were as follows (Table 2):
  a. 24 hours postoperatively, CNB (6.4, 3.29) vs SNB + IV Lido (8, 2.29)
  b. 48 hours, CNB (2.4, 5.92) vs SNB + IV Lido (4, 2.00)
  c. 72 hours, CNB (2, 2.82) vs SNB + IV Lido (2.8, 3.12)
3. **Non-opioids analgesics** There were no statistical differences among groups. Total non-narcotic analgesic usage was as follows (Table 3):
  a. Acetaminophen (0-72 hours): acetaminophen was given at scheduled dosing periods for a maximum of 72 hours, CNB (3000, 136.57) vs SNB + IV Lido (3000, 195.29)
  b. Dexmedetomidine (0-24 hours): dexmedetomidine was infused for a maximum of 24 hours postoperatively while the patient was in the ICU following VATS, CNB (181.90, 43.59) vs SNB + IV Lido (99.99, 57.92).
  c. Ketamine (0-72 hours): ketamine was infused postoperatively for a maximum of 72 hours postoperatively, CNB (87.82, 8.51) vs SNB + IV Lido (78.53, 19.41)
  d. Ketorolac (0-72 hours): ketorolac was given at scheduled dosing periods for a maximum of 72 hours, CNB (30.00, 4.39) vs SNB + IV Lido (30.00, 6.07)

**Table 2.** Comparison of pain scores and opiate consumption (MIVE) between groups (median, CI)

|  | **Pain**<br>**Scores 24**<br>**hrs** | **Pain**<br>**Scores**<br>**48 hrs** | **Pain**<br>**Scores**<br>**72 hrs** | **MIVE 0-**<br>**24 hrs** | **MIVE 24-**<br>**48 hrs** | **MIVE 48-**<br>**72 hrs** |
| --- | --- | --- | --- | --- | --- | --- |
| **CESPB, SESPb,**<br>**CQLB, SQLB** |  |  |  |  |  |  |
| Count | 105 | 96 | 73 | 105 | 96 | 73 |
| Median (CI 95%) | 5 (0.45) | 4 (0.51) | 4 (0.52) | 7.2 (2.19) | 3.6 (3.75) | 2 (2.09) |
| **CESPB & CQLB** |  |  |  |  |  |  |
| Count | 65 | 60 | 47 | 65 | 60 | 47 |
| Median (CI 95%) | 5 (0.59) | 4 (0.61) | 4 (0.64) | 6.4 (3.29) | 2.4 (5.92) | 2 (2.82) |
| **SESPB & SQLB +**<br>**IV Lido** |  |  |  |  |  |  |
| Count | 40 | 36 | 26 | 40 | 36 | 26 |
| Median (CI 95%) | 5 (0.75) | 4 (0.94) | 4 (0.94) | 8 (2.29) | 4 (2.00) | 2.8 (3.12) |
| **CESPB (Thoracic)** |  |  |  |  |  |  |
| Count | 34 | 30 | 21 | 34 | 30 | 21 |
| Median (CI 95%) | 5 (0.85) | 4.5<br>(0.84) | 5 (1.21) | 4.4 (5.47) | 1 (2.16) | 2 (2.46) |
| **SESPB + IV Lido**<br>**(Thoracic)** |  |  |  |  |  |  |
| Count | 17 | 13 | 8 | 17 | 13 | 8 |
| Median (CI 95%) | 4 (1.01) | 4 (2.05) | 4 (2.31) | 7.6 (2.52) | 4 (2.46) | 4.2 (7.74) |
| **CQLB (Abdominal)** |  |  |  |  |  |  |
| Count | 31 | 30 | 26 | 31 | 30 | 26 |
| Median (CI 95%) | 5 (0.85) | 4 (0.93) | 4 (0.80) | 9.2 (3.72) | 7.6 (11.72) | 4 (4.77) |
| **SQLB + IV Lido**<br>**(Abdominal)** |  |  |  |  |  |  |
| Count | 23 | 23 | 18 | 23 | 23 | 18 |
| Median (CI 95%) | 5 (1.13) | 4 (1.07) | 4 (1.09) | 8.8 (3.63) | 2.4 (2.96) | 2 (3.57) |

Abbreviations: CESPB, continuous Erector Spinae block; SESPb, single shot Erector Spinae block; CQLB, continuous Quadratus Lumborum block; SQLB, single shot continuous Quadratus Lumborum block; IV Lido, Intravenous Lidocaine infusion; CNB, continuous nerve block; SNB, single shot nerve block. Overall Data represented by CESPB, SESPb, CQLB, SQLB. CNBs represented by CESPB & CQLB. SNBs represented by SESPb + IV Lido. Thoracic represents VATS surgery. Abdominal represents major abdominal surgery.

**Table 3.**
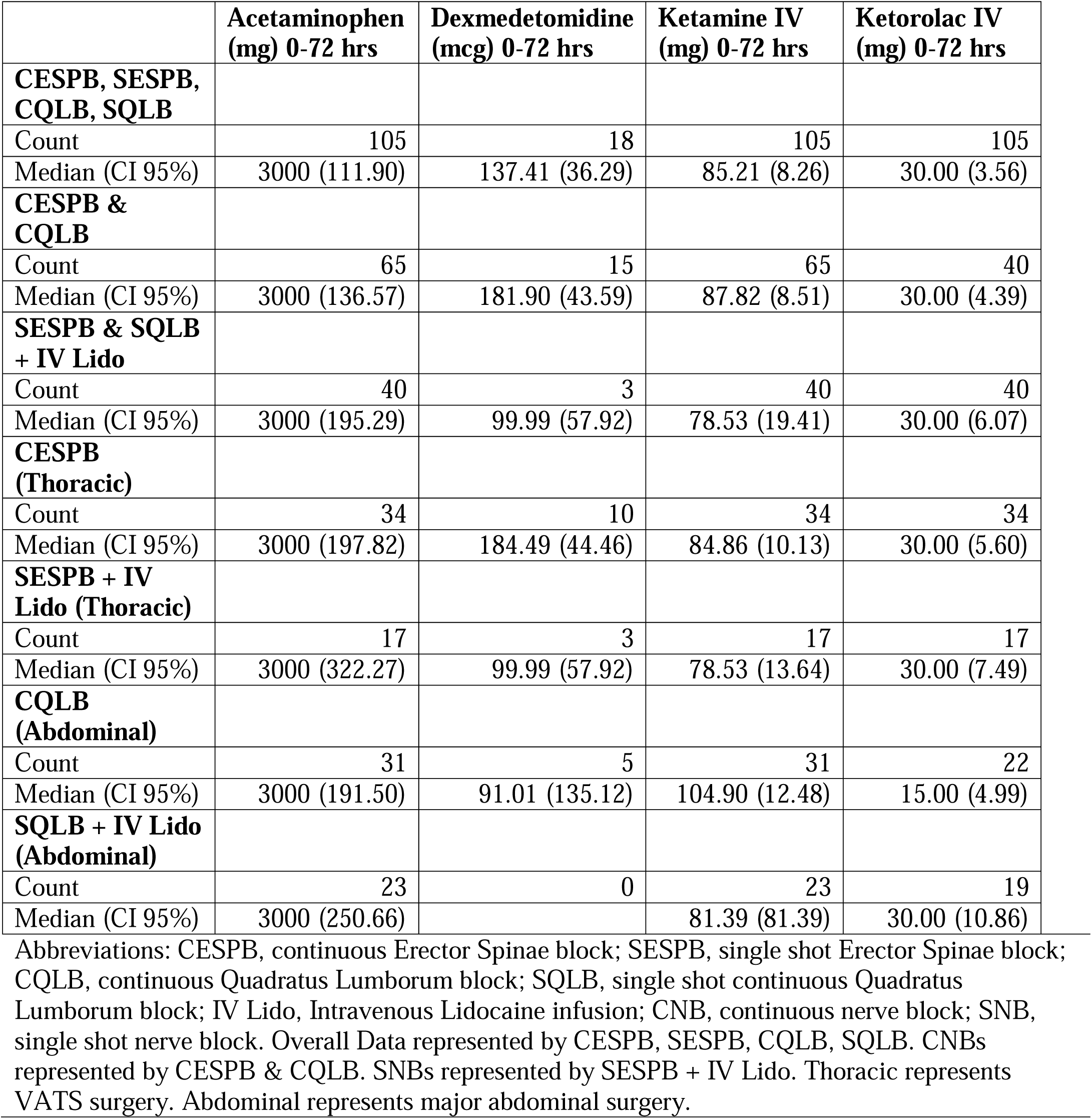
Comparison of non-opiate analgesics usage between groups (median, CI)

### Thoracic Surgery – CESPB vs SESPB + IV Lido

1. **Pain scores** There were no statistical differences among groups. VAS scores were as follows (Table 2):
  a. 24 hours postoperatively, CNB (5, 0.85) vs SNB + IV Lido (4, 1.01)
  b. 48 hours, CBN (4.5, 0.84) vs SNB + IV Lido (4, 2.05)
  c. 72 hours, CNB (5, 1.12) vs SNB + IV Lido (4, 2.31)
2. **Opioid consumption** There were no statistical differences among groups. MIE were as follows (Table 2):
  a. 24 hours postoperatively, CNB (4.4, 5.47) vs SNB + IV Lido (7.6, 2.52)
  b. 48 hours, CNB (1, 2.16) vs SNB + IV Lido (4, 2.46)
  c. 72 hours, CNB (2, 2.46) vs SNB + IV Lido (4.2, 7.74)
3. **Non-opioids analgesics** There were no statistical differences among groups. Total non-narcotic analgesic usage was as follows (Table 3):
  a. Acetaminophen (0-72 hours): acetaminophen was given at scheduled dosing periods for max 72 hours, CNB (3000, 197.82) vs SNB + IV Lido (3000, 322.27)
  b. Dexmedetomidine (0-24 hours): dexmedetomidine was infused for a maximum of 24 hours postoperatively while the patient was in the ICU following VATS, CNB (184.49, 44.46) vs SNB + IV Lido (99.99, 57.92)
  c. Ketamine (0-72 hours): ketamine was infused postoperatively for a maximum of 72 hours postoperatively, CNB (84.86, 10.13) vs SNB + IV Lido (78.53, 13.64)
  d. Ketorolac (0-72 hours): ketorolac was given at scheduled dosing periods for a maximum of 72 hours, CNB (30, 5.60) vs SNB + IV Lido (30, 7.49)

### Abdominal Surgery – CQLB vs SQLB + IV Lido

1. **Pain scores** There were no statistical differences among groups. OPS were as follows (Table 2):
  a. 24 hours postoperatively, CNB (5, 0.85) vs SNB + IV Lido (5, 1.13)
  b. 48 hours, CNB (4, 0.93) vs SNB + IV Lido (4, 1.07)
  c. 72 hours, CNB (4, 0.80) vs SNB + IV Lido (4, 1.09)
2. **Opioid consumption** There were no statistical differences among groups. MIE were as follows (Table 2):
  a. 24 hours postoperatively, CNB (9.2, 3.72) vs SNB + IV Lido (8.8, 3.63)
  b. 48 hours, CNB (7.6, 11.72) vs SNB + IV Lido (2.4, 2.96)
  c. 72 hours, CNB (4, 4.77) vs SNB + IV Lido (2, 3.57)
3. **Non-opioids analgesics** There were no statistical differences among groups. Total non-narcotic analgesic usage was as follows (Table 3):
  a. Acetaminophen (0-72 hours): acetaminophen was given at scheduled dosing periods for a maximum of 72 hours, CNB (3000, 191.50) vs SNB + IV Lido (3000, 250.66)
  b. Dexmedetomidine (0-24 hours): dexmedetomidine was infused for a maximum of 24 hours postoperatively while the patient was in the ICU following VATS, CNB (91.01, 135.12) vs SNB + IV Lido (0)
  c. Ketamine (0-72 hours): ketamine was infused postoperatively for a maximum of 72 hours postoperatively, CNB (104.90, 12.48) vs SNB + IV Lido (81.39, 81.39)
  d. Ketorolac (0-72 hours): ketorolac was given at scheduled dosing periods for a maximum of 72 hours, CNB (15, 4.99) vs SNB + IV Lido (30, 10.86)

## Discussion

The use of intravenous lidocaine infusions for analgesia has been advocated since 1951. ^39^ Although, it is possible to find studies published on the topic in the 80s, the recent development of laparoscopic surgery has led to a renewed interest in the technique. The average dose of lidocaine reported in the case of an infusion of 24hrs and 48hrs is 2,894 mg and 4,678 mg, respectively. Cassuto et al, 1985 ^40^ advocated the use of 3,460 mg for 24 hrs in an article titled “Inhibition of postoperative pain by continuous low dose intravenous infusion of lidocaine”. Our protocol is based on the use of 2,400 mg, which is less than most authors reported for both 24 and 48 hrs of infusions. The chosen dose is based on our previous experience with the use of the same protocol in patients undergoing procedures such as lung wedge and abdominal surgery.

Our data support that, in the context of a multimodal approach to perioperative pain following either VATS or major open abdominal surgery, the use of a single injection nerve block followed by a continuous infusion of IV lidocaine at 50 mg/hr provides similar perioperative analgesia than a continuous ESP or QL plane block in patients undergoing VATS and major abdominal surgery, respectively.

Recent reviews provide evidence supporting the use of continuous intravenous infusions of lidocaine for postoperative analgesia following several different types of surgery, including abdominal, VATS, and spine.^31-40^ Our study demonstrated that a continuous intravenous infusion of lidocaine started in the recovery room after a single injection nerve block performed prior to surgery is as effective as a continuous nerve block. Both techniques include a preoperative injection of local anesthetic into a fascial plane that is followed by a continuous local anesthetic infusion started in the recovery room, administered either intravenously or through the nerve block catheter.

Although only a limited number of studies have compared facial plane nerve blocks to more established approaches, such as paravertebral blocks and/or lumbar plexus blocks, the use of ultrasound-guided fascial plane blocks, including ESP and QL blocks, have gained popularity. Fascial plane blocks offer the advantage of reduced risks for complications, such as pneumothorax and bleeding. The associated risk of major bleeding is reduced, as local anesthetic is injected into a fascial plane, far away from the major vessels that travel near large nerves. The risk of pneumothorax is reduced, as the ESP is found superficial to the transverse process, which is easily identified and located centimeters superficial to the pleura. When comparing single injection nerve blocks to continuous nerve blocks, there is no risk of intravascular catheter migration in a single injection nerve block, as there is no catheter. This makes our model not only relevant but practical as well.

Lidocaine’s half-life is estimated to be between 90-120 min. In the absence of an initial bolus, reaching steady state requires between 7.5 and 12 hours of infusion. The administration of an initial bolus, although of theoretical value to reach steady state more rapidly, would not be of value in the context of surgery, as it would require cardiac monitoring, which is not always available prior to surgery. Under these conditions, the performance of a single injection nerve block allows for the provision of an analgesic bridge prior to a lidocaine infusion reaching a steady state concentration.

Evidence supports the concept that preemptive analgesia can significantly reduce postoperative pain and opioid consumption when compared to beginning pain management either intra-operatively or postoperatively. This is best achieved by performing a regional never block prior to surgery. Unfortunately, the skills and resources required to perform a nerve block are a finite entity. A single injection nerve block requires much less time and resources when compared to a continuous technique. SNBs have several advantages when compared to CNBs: their performance requires less time, there is a lower risk of bleeding or infections, there is no risk of perineural catheter dislodgment or migration, and they require a lower overall cost.

Although the main mechanism of action of lidocaine is blocking voltage-gated sodium channels (VGSC/NaVs), it has been demonstrated that lidocaine can also reduce the peak currents of sodium channels and accelerate the deactivation process to reduce the excitability of sensory fibers, leading to a reduction of pain perception.^24^ At the level of the central nervous system, lidocaine has been reported to control the presynaptic release of glutamate from presynaptic terminals of the spinal substantia gelatinosa neuron. It has also been reported to interfere with the G-couple proteins of the GABA, substance P, and neurokinin-1 receptor.^39^

Our findings need to be confirmed using a prospective randomized design, which is currently in progress, but our data are especially relevant in modern healthcare systems where increasing emphasis is placed upon quality, efficiency, and appropriate utilization of resources.

The limitation of this study includes the fact it is a retrospective analysis. However, in each case, the surgery was performed under the same ERAS protocol. Additionally, patients were not randomized to each treatment arm, which can only be achieved using a prospective, randomized design.

## Conclusion

This retrospective analysis suggests that a single injection nerve block followed by a continuous IV lidocaine infusion may be as effective as a continuous nerve block in patients undergoing major abdominal surgery and VATS. A prospective randomized study is required to confirm these findings.

## Data Availability

All data produced in the present study are available upon reasonable request to the authors.

## Acknowledgments

None.

